# The impact of COVID-19 on African American communities in the United States

**DOI:** 10.1101/2020.05.15.20096552

**Authors:** Elena Cyrus, Rachel Clarke, Dexter Hadley, Zoran Bursac, Mary Jo Trepka, Jessy G. Dévieux, Ulas Bagci, Debra Furr-Holden, Makella Coudray, Yandra Mariano, Sandra Kiplagat, Ines Noel, Gira Ravelo, Michelle Paley, Eric F. Wagner

## Abstract

**Importance:** The novel Coronavirus Disease 2019 (COVID-19), declared a pandemic in March 2020, may present with disproportionately higher rates in underrepresented racial/ethnic minority populations in the United States, including African American communities who have traditionally been over-represented in negative health outcomes.

**Study Objective:** To understand the impact of the density of African American communities (defined as the percentage of African Americans in a county) on COVID-19 prevalence and death rate within the three most populous counties in each U.S. state and territory (n=152).

**Design:** An ecological study using linear regression was employed for the study.

**Setting:** The top three most populous counties of each U.S. state and territory were included in analyses for a final sample size of n=152 counties.

**Participants:** Confirmed COVID-19 cases and deaths that were accumulated between January 22, 2020 and April 12, 2020 in each of the three most populous counties in each U.S. state and territory were included.

**Main outcome measures:** Linear regression was used to determine the association between African American density and COVID-19 prevalence (defined as the percentage of cases for the county population), and death rate (defined as number of deaths per 100,000 population). The models were adjusted for median age and poverty.

**Results:** There was a direct association between African American density and COVID-19 prevalence; COVID-19 prevalence increased 5% for every 1% increase in county AA density (p<.01). There was also an association between county AA density and COVID-19 deaths, such; the death rate increased 2 per 100,000 for every percentage increase in county AA density (p=.02).

**Conclusion:** These study findings indicate that communities with a high African American density have been disproportionately burdened with COVID-19. Further study is needed to indicate if this burden is related to environmental factors or individual factors such as types of employment or comorbidities that members of these community have.

**Key Points:** *Question:* What is the impact of the density of African American communities (defined as the percentage of African Americans in a county) on COVID-19 prevalence and death rate by U.S. counties.

*Findings:* This ecological study including the three most populous counties in each U.S. state and territory, found that COVID-19 prevalence increased 5% for every percentage increase in county African American density, and that the death rate increased 2 per hundred thousand for every percentage increase in county African American density. Both were statistically significant increases.

*Meaning:* These study findings indicate that communities with a high African American density have been disproportionately burdened with COVID-19.

## Background

The Coronavirus Disease (COVID-19), was first reported in December of 2019 in Wuhan, China and by January 31, 2020, the World Health Organization declared the outbreak to be a Public Health Emergency of International concern ^1^ (see Figure 1 for the timeline). As of April 18, 2020 in the United States (U.S.), there were approximately 691,000 COVID-19 cases and 35,000 COVID-19 related deaths ^2^. As of April 20, 2020 in the United States (U.S.), there were approximately 773,863 COVID-19 cases and 41,400 COVID-19 related deaths ^2^. As of April 28, 2020 in the United States (U.S.), there were approximately 982,000 COVID-19 cases and 56,000 COVID-19 related deaths. ^2^ As data are collected and analyzed, researchers can better understand risk factors associated with disease transmission.

**Figure 1:**
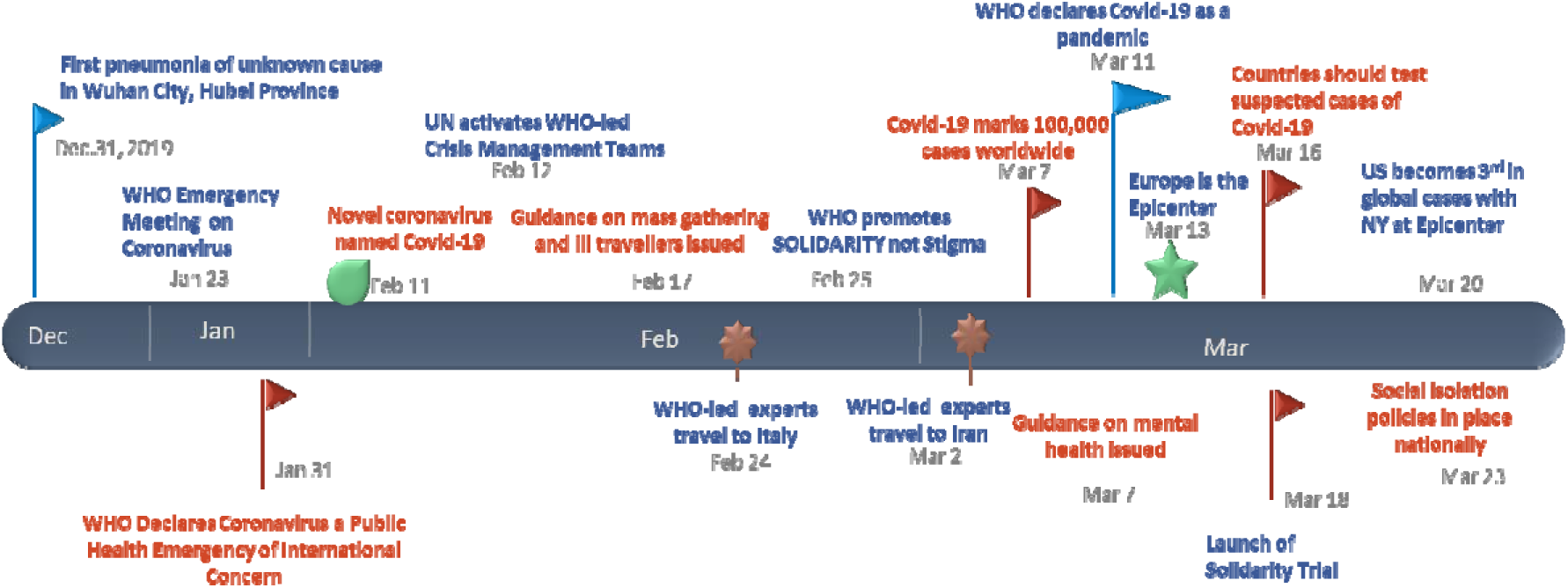
Timeline of COVID-19 pandemic events to from December 2019 to March 2020.

Thus far, one of the findings of ongoing and evolving research related to social disparities and health inequities in the U.S. is that, compared to other races/ethnicities, Blacks/ African Americans are disproportionately represented in the COVID-19 epidemic. ^3,4^ This is not a unique experience for African American communities, as prevailing research provide evidence of Blacks experiencing overall higher morbidities, earlier onset of morbidities and higher mortality rates when compared to non-Hispanic Whites in the U.S.^5,6^ Specifically, African Americans adults a) are 60% more likely to have diabetes than non-Hispanic White adults;^7^ b) are responsible for 42% of new HIV cases even though they make up only 13% of the population;^8^ c) are 20% more likely to die from heart disease when compared to non-Hispanic Whites;^9^ d) have a higher prevalence of asthma;^10^ and e) are 1.3 times more likely to be obese than non-Hispanic Whites.^11^

Social determinants of health such as the places individuals live and work, their access to quality health care, and the resources to lead a healthy lifestyle play a major role in determining health status and health outcomes.^5,12^ Additionally, a higher percentage of Blacks live in densely populated communities and work as essential workers in service industries where they are at greater risk of exposure.^13,14^ Public health research shows that in comparison to their counterparts, Blacks are more likely to be uninsured or underinsured and as a result have lower access to quality healthcare and tend to receive lower quality healthcare.^5,15^

Similar to influenza, for COVID-19, vulnerable and marginalized populations may be at greater risk for having more severe outcomes or death if they contract the virus. For COVID-19, vulnerable and at-risk populations (i.e. those with comorbidities like asthma, diabetes, serious heart conditions, chronic kidney disease and severe obesity) are at greater risk of negative outcomes.^16^ Due to many social and economic factors, Blacks are disproportionately affected by a number of these conditions ^6,17^ and are more likely to have poor outcomes if COVID-19 is contracted. This preliminarily study aims to understand the relationship between AA density and other social determinants with COVID-19 prevalence and death rates nationally.

## Methods

An ecological analysis was completed using multivariable linear regression. The three most populous counties of each U.S. state and territory were identified and included in the data analysis, with the exception of Washington, DC which is not considered a state so had only one entry, and California for which four counties and parishes were included (see Appendix A for the full list). This resulted in a final sample size of 152 counties and parishes. Data were analyzed for cases collected between January 22, 2020 and April 12, 2020. The data were sourced from USA Facts,^18^ and population estimates were derived from the U.S. and population estimates were derived from the U.S. Census.^19^

Independent variables were African American density (percentage of county/parish population who identified as African American), poverty level (percent of county/parish population at the defined poverty level), and median age for the counties/parishes. Primary outcomes were the percentage of COVID-19 confirmed cases (prevalence) and rate of COVID-19 deaths per 100,000 persons/parish (death rate). Prevalence was used as estimate of the existing burden of disease by county, and death rate provides inference on the health status of a community and the overall status of a public health system.^20^

Descriptive statistics including overlapping frequencies were used to describe COVID-19 prevalence and African American density by county. Linear regression models were used to determine the relationship between AA density and poverty, and, COVID-19 prevalence and death rates across U.S. counties and parishes. Both models for prevalence and death rate were adjusted for median age. All results were considered to be statistically significant at an alpha level of 0.05. Analyses were conducted using SPSS version 22.0.^21^

## Results

Two multivariable linear regression models were run to predict two health outcomes - confirmed COVID-19 cases or prevalence, and death rate.

### Prevalence

A multivariable linear regression was conducted to predict COVID-19 prevalence based on AA density and county/parish poverty level. There was a significant regression equation for this model. (F (3,148) = 5.7, *p*<0.01) with an R^2^ of 0.32. After adjusting for the county median age, there was a direct association between county AA ethnic density and COVID-19 prevalence; COVID-19 prevalence increased 5% for 1% increase in county AA density (*p*<.01). There was no significant association between county poverty level and COVID-19 cases.

### Death rate

A multivariable linear regression was conducted to predict COVID-19 death rate based on AA density and county/parish poverty level. There was a significant regression equation for this model. (F (3,148) = 5.15, *p*=.02) with an R^2^ of 0.30. After adjusting for the county median age, there was an association between county AA ethnic density and COVID-19 deaths; the death rate increased 2 per 100,000 for every percentage point increase in county AA density (*p*=.02). Although not statistically significant, the data trended toward an association between county poverty level and the death rate (p=0.12). (See Table 1 for all results)

**Table 1:**
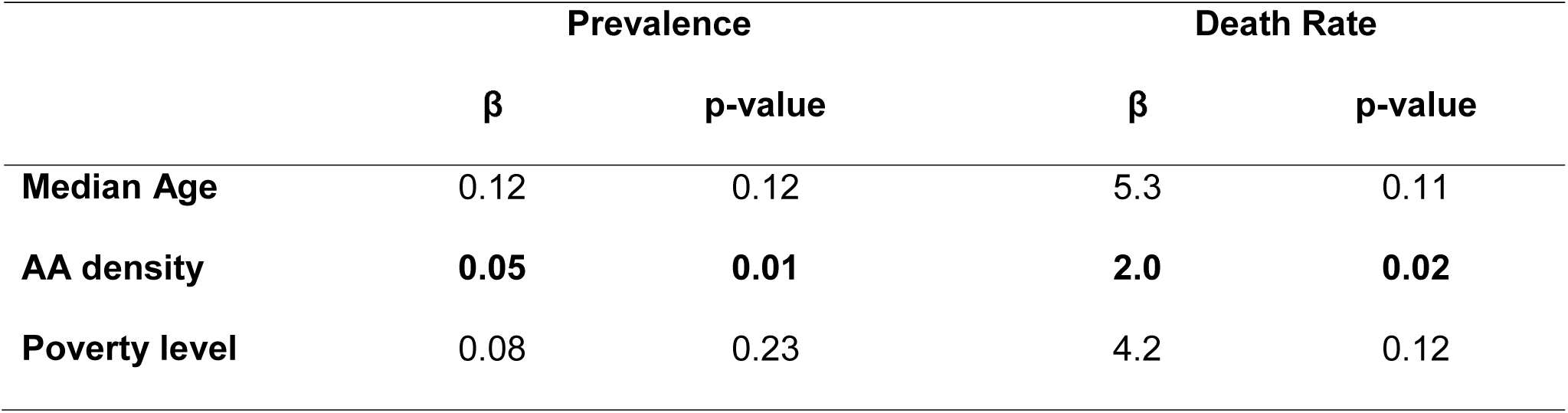
Linear Regression for AA density and poverty level on COVID-19 prevalence and death rate US counties/parishes, January 22 to April 12, 2020 (n=152)

Illustration of the descriptive analysis (Figure 2) demonstrates in some geographical areas or counties with higher AA density, there are peaks in prevalence.

**Figure 2:**
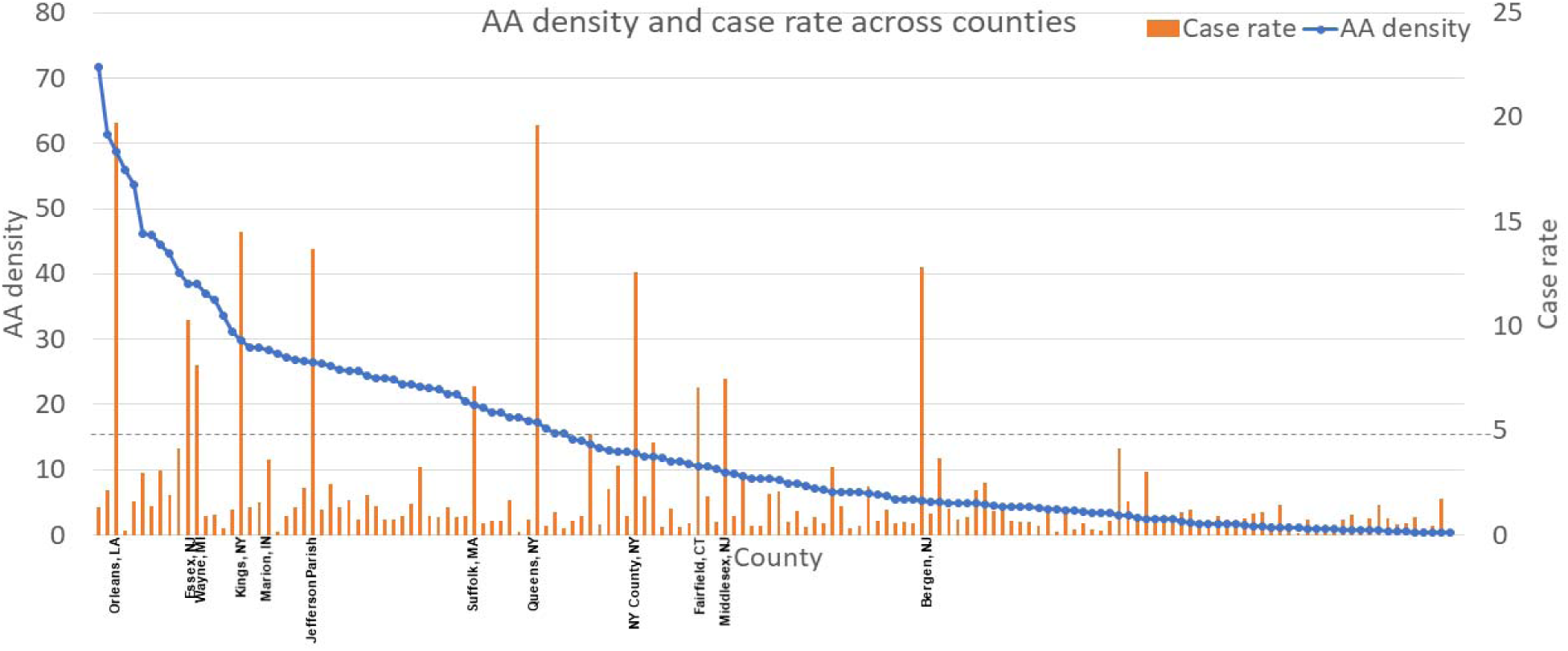
Covid-19 prevalence by 152 counties/parishes in the United States January 22 to April 12, 2020 (n=152)

## Discussion

According to this ecologic study, in the U.S., higher African American density was more strongly associated with COVID-19 prevalence and death than higher median age in a community. These study findings support earlier research that suggested differences in cases, hospitalizations and deaths could be due to the high prevalence of comorbidities among African Americans and the jobs as essential that many African Americans do, placing them at a higher risk of exposure to the virus.^3,4^

Most U.S. states and territories have peaked in incidence and have started to see a decline in new cases.^22^ Despite this general abatement, some researchers have predicted a resurgence of cases in the later part of year due to several factors including relaxation of policies related to mitigation and social distancing, and possibly also an increase in travel related/imported cases due to less travel restrictions.^23^

In the anticipated surge in the later part of the year, the most at-risk vulnerable populations, including African American populations, will be disproportionately represented, more so than in the first wave of COVID-19 infections. Much of this may be due to the living conditions of many African Americans, where the close proximity and housing conditions result in a higher likelihood of community spread.^13^ Additionally, it is more difficult for African Americans to be tested for COVID-19, and if they are able to receive the test and are tested positive, they are less likely to receive quality care.^24^

In the long term, with the availability of vaccines and efficacious therapeutic lines,^25^ COVID-19 will likely become endemic, where it will constantly exist amongst minority populations,^26^ as is the other case with other historical pandemics.^27^ Part of working toward this ‘new normal’ successfully and living with this novel coronavirus is to use all scientific and research efforts to minimize the number of fatalities. Implicit in this approach is ensuring that there is adequate testing in underserved areas such as AA communities.^28,29^ Screening and services should be comprehensively available regardless of insurance status or ability of the individuals to pay for medical care. Widespread availability of these services as well as vaccines and treatment when they are available, will contribute to the reduction in overall incidence, transmission and community spread.^29,30^

## Limitations

There are several limitations associated with this study, First, this is an ecologic analysis and there is a lot of potential unmeasured confounding, Second, non-Hispanic Whites were not included in the analysis as a possible reference group. A third limitation was that we did not account for testing differences between the counties. Finally, it is possible that prevalence estimates can be skewed depending on the availability of the test per county population.

## Conclusion

Study findings indicate an association between communities that have higher percentage of African Americans and negative COVID-19 health outcomes including higher prevalence and higher death rates. Additionally, although not statistically significant in this study, the data suggested that the odds of surviving the epidemic may also be related to poverty levels suggesting that other at-risk minority populations, affected by poverty, may also be disproportionately affected.^31^ Further comprehensive analysis is needed to understand state, community and individual levels of social determinants on COVID-19 health outcomes for all racial/ethnic minority and other vulnerable populations living in the United States.

## Data Availability

The data that support the findings of this study are openly available in USA Facts at https://usafacts.org/visualizations/coronavirus-covid-19-spread-map/ and U.S. Census at census.gov. (reference number: 18, 19).

https://www.census.gov

## Acknowledgements

The project was supported by Award Number from the National Institute on Drug Abuse (NIDA) at the National Institutes of Health and Award K99DA046311; NIMHD 1 S21 MD010683-01; NIMHD U54MD002266; NIMHD U54MD012393 and NIAAA U34AA026219. The content is solely the responsibility of the authors and does not necessarily represent the official views of the National Institutes of Health.

